# The Role of Neuroimaging in Evolving TBI Research and Clinical Practice

**DOI:** 10.1101/2023.02.24.23286258

**Authors:** Emily L Dennis, Finian Keleher, David F Tate, Elisabeth A Wilde

## Abstract

Neuroimaging technologies such as computed tomography (CT) and magnetic resonance imaging (MRI) have been widely adopted in the clinical diagnosis and management of traumatic brain injury (TBI), particularly at the more acute and severe levels of injury. Additionally, a number of advanced applications of MRI have been employed in TBI-related clinical research with great promise, and researchers have used these techniques to better understand underlying mechanisms, progression of secondary injury and tissue perturbation over time, and relation of focal and diffuse injury to later outcome. However, the acquisition and analysis time, the cost of these and other imaging modalities, and the need for specialized expertise have represented historical barriers in extending these tools in clinical practice. While group studies are important in detecting patterns, heterogeneity among patient presentation and limited sample sizes from which to compare individual level data to well-developed normative data have also played a role in the limited translatability of imaging to wider clinical application. Fortunately, the field of TBI has benefitted from increased public and scientific awareness of the prevalence and impact of TBI, particularly in head injury related to recent military conflicts and sport-related concussion. This awareness parallels an increase in federal funding in the United States and other countries allocated to investigation in these areas. In this article we summarize funding and publication trends since the mainstream adoption of imaging in TBI to elucidate evolving trends and priorities in the application of different techniques and patient populations. We also review recent and ongoing efforts to advance the field through promoting reproducibility, data sharing, big data analytic methods, and team science. Finally, we discuss international collaborative efforts to combine and harmonize neuroimaging, cognitive, and clinical data, both prospectively and retrospectively. Each of these represent unique, but related, efforts that facilitate closing gaps between the use of advanced imaging solely as a research tool and the use of it in clinical diagnosis, prognosis, and treatment planning and monitoring.

## Data Collection for Publication and Funding Trends

To understand publication trends, we searched PubMed for publications focused on neuroimaging in human patients with TBI, concussion, or subconcussive impact exposure. Search terms are detailed in **Supplementary Note 1**. Resulting lists were reviewed by ELD and FK to remove papers that were out of scope, and to classify the remaining papers based on the imaging modality/ies used and the patient population(s) studied. These designations were made based on the paper title, abstract, or full paper if neither the title nor abstract provided enough detail. Individual case reports and postmortem reports were excluded. Papers that could not be classified by the title or abstract and were behind a paywall for the authors (who had institutional access through the University of Utah) were also excluded. The patient population categories included the following: moderate/severe, mild civilian, military, sports, pediatric, geriatric, coma/vegetative state, penetrating TBI, or intimate partner violence/domestic violence (IPV/DV). If papers on mild TBI (mTBI) specified the injury mechanism as sports-related or military-relevant, they were categorized as such and not double-counted as civilian mTBI. The imaging modalities were: T1-weighted MRI, T2-weighted MRI, diffusion MRI (dMRI, including diffusion tensor, diffusion kurtosis, diffusion weighted, and high angular resolution diffusion imaging [HARDI]), functional MRI (fMRI), resting state fMRI (rsfMRI), magnetic resonance spectroscopy (MRS), positron emission tomography (PET), single photon emission computed tomography (SPECT), electroencephalography (EEG), magnetoencephalography (MEG), arterial spin labeling (ASL), quantitative susceptibility mapping/susceptibility weighted imaging (QSM/SWI), near-infrared spectroscopy (NIRS, including fNIRS), and other. Computed tomography (CT) was not included. All years were queried up to the search date (July 29, 2022), with the first paper identified in 1966. Papers that were not available on PubMed by the search date were not included.

To detail funding trends from the National Institutes of Health (NIH) in traumatic brain injury and concussion where imaging was a specific focus of the objectives, NIH RePORTER was searched, search terms detailed in **Supplementary Note 2**. The search terms to query Congressionally Directed Medical Research Programs (CDMRP)) records are listed in **Supplementary Note 3**. Funding records for the Veterans Administration were listed in NIH RePORTER but without funding amounts, so the number of projects was summarized rather than funding amounts. All available fiscal years (FY) were queried (NIH RePORTER has records beginning in 1985, CDMRP beginning in 2009). The NIDILRR (National Institute on Disability, Independent Living and Rehabilitation Research) program database was also searched using the same search terms. The list of projects was reviewed by ELD and FK to remove projects that were out of scope, and to classify the remaining projects based on the patient population(s) studied from the titles and/or project abstracts.

### Publication Trends

The publication data show an accelerating increase in papers applying neuroimaging techniques to TBI. The publication rate rises steeply in 2010, with an increase of around 250% over the last decade. The current year (2022) is included in the graphs but only represents a partial year. Examination of publication rates classified by patient population (**Figure 1**) indicates that the most pronounced increase appears to be in mTBI, including sports-related head injuries and military-relevant TBI. The increase in publication of papers with a focus on pediatric TBI papers is relatively consistent over the years. The fewest papers have focused on penetrating TBI and coma/vegetative state, which is to be expected given that the prevalence of these more severe forms of TBI represent a very small proportion of injuries annually (Leo and McCrea, 2015; Løvstad et al., 2014). IPV-related TBI also has very few publications, which is also anticipated given the challenges in studying this population. Lastly, there are few papers on TBI in older adults.

**Figure 1.**
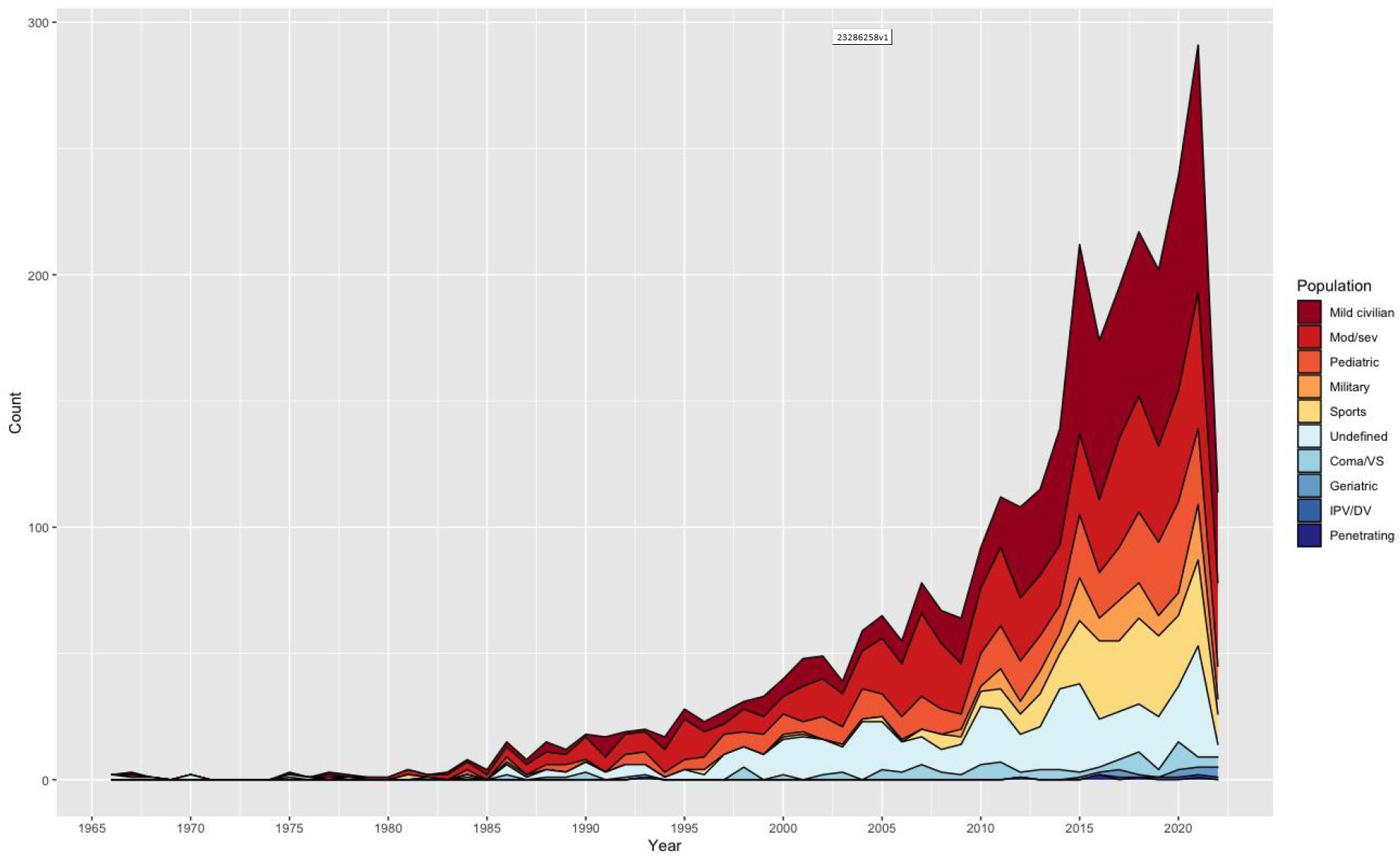
Publication trends classified by patient population. VS=vegetative state, IPV/DV=intimate partner violence/domestic violence, mod/sev=moderate/severe.

Examination of the publication data organized by neuroimaging modality (**Figure 2**) reveals a higher publication rate of data using T1-weighted MRI as well as a consistent increase in the use of this sequence over time. Imaging techniques such as EEG, PET and MRS show a more consistent rise over time. The pace of publication using fMRI, dMRI, or multimodal approaches has increased in the last decade. SPECT, MEG, ASL, and NIRS are the least published, although MEG and ASL show signs of accelerated adoption, while SPECT has a fairly consistent rate of usage over the past two decades. The two newest modalities are rsfMRI and QSM/SWI, both of which show increasing adoption.Publication trends with scientific papers separated from review/editorial/ commentary papers are shown in **Supplementary Figures 1-3**.

**Figure 2.**
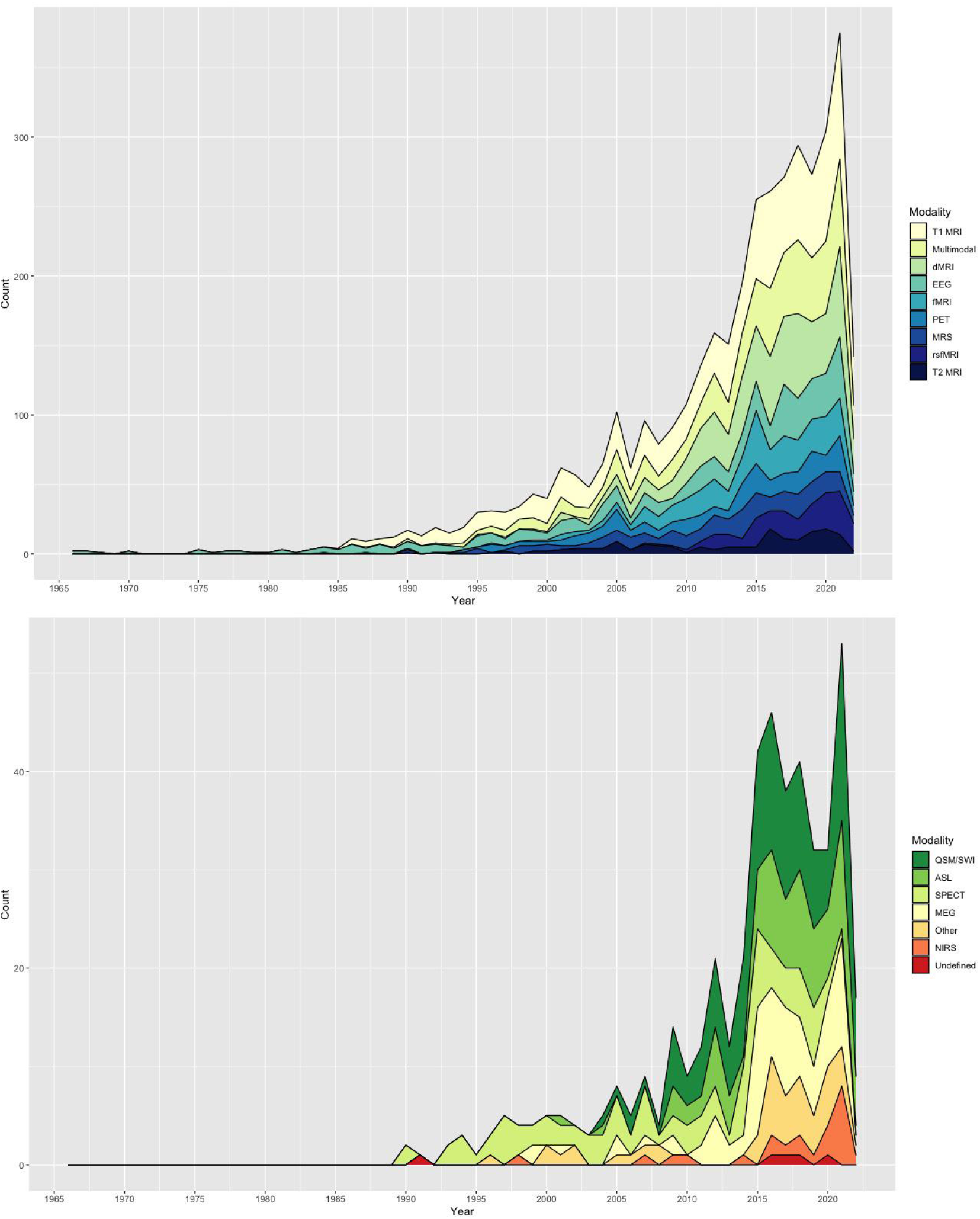
Publication trends classified by modality. Data are split into two panels for easier viewing. dMRI=diffusion magnetic resonance imaging, EEG=electroencephalography, fMRI=functional MRI, MRS=magnetic resonance spectroscopy, PET=positron emission tomography, rsfMRI=resting state fMRI, ASL=arterial spin labeling, MEG=magnetoencephalography, NIRS=near-infrared spectroscopy, QSM=quantitative susceptibility mapping, SWI=susceptibility weighted imaging, SPECT=single photon emission computed tomography.

## Funding Trends

Federal funding trends (United States) in imaging in TBI generally parallel publication trends, which also demonstrate dramatic increases over the past two decades (**Figure 3**). With regard to funding by the NIH, there was clearly a large increase in funding in fiscal year 2014. This may be due primarily to an increase in the number of funded projects (from 19 to 31). Around this time (2013-2017), there were also a number of large, multi-site U01 projects funded by the NINDS, including the Late Effects of TBI (LETBI) (Dams-O’Connor et al., 2013) and Transforming Research and Clinical Knowledge of TBI (TRACK-TBI) (Yue et al., 2013). It is notable that this increase occurred despite an overall decrease in the NIH total budget (in adjusted dollars). This increase in funding is apparent across patient populations in TBI, but the growth is again particularly evident in studies with a focus on mild TBI, including sports-related head injuries and military-relevant TBI. The apparent decrease in funding for fiscal year 2022 is related to the use of a partial year of data which was available at the time of data analysis rather than a true decrease. The apparent drop in funding for fiscal year 2018 partly reflects the end of the funding cycle for two large projects ending (LETBI and TRACK-TBI) and may also be influenced by no-cost extensions (NCEs).

**Figure 3.**
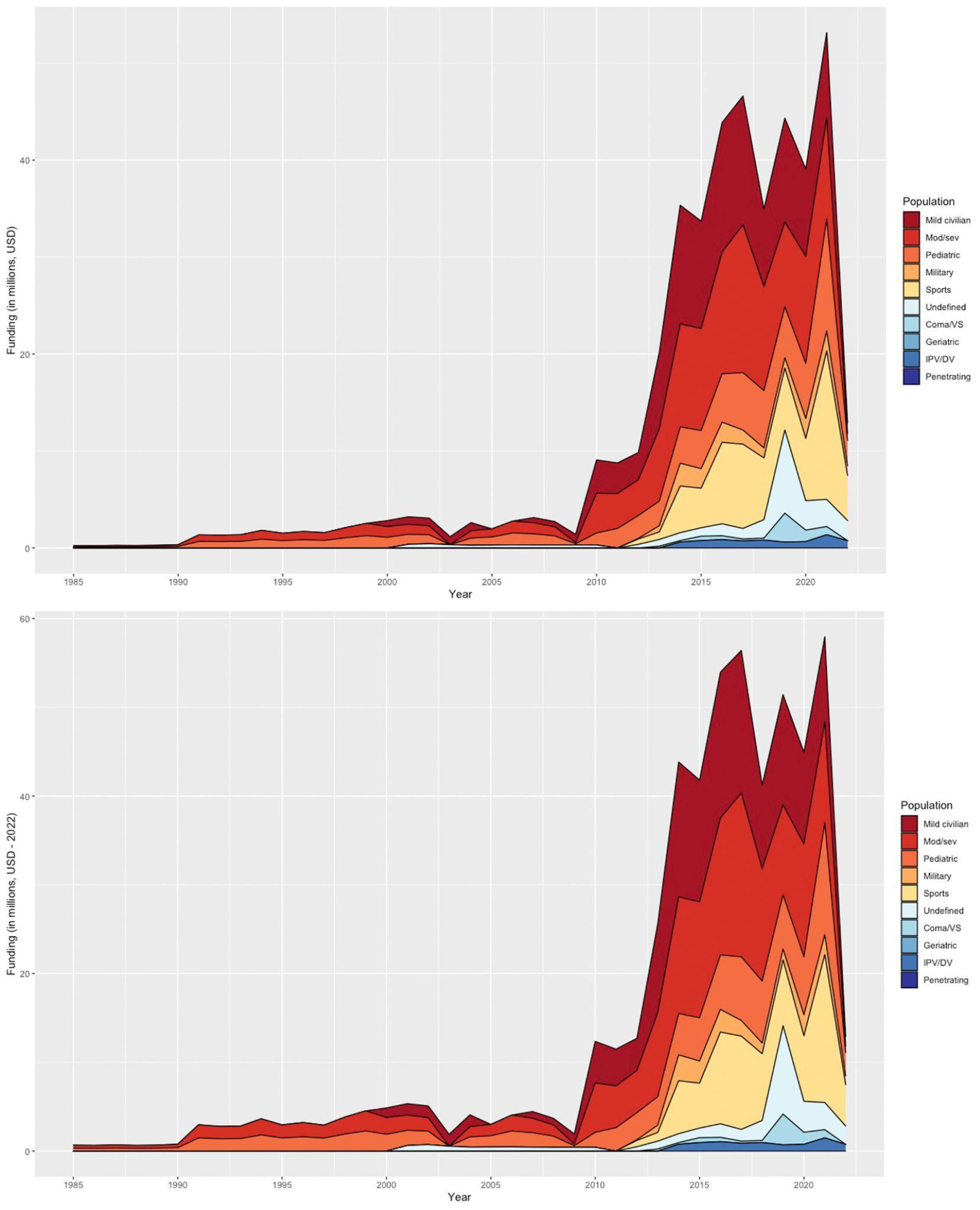
NIH funding trends classified by population. Amounts are shown in millions, in US Dollars. VS=vegetative state, mod/sev=moderate/severe, IPV/DV=intimate partner violence/ domestic violence. During this time period, there were no projects identified with geriatric TBI or penetrating TBI as the main focus. Records dating back to 1985 were queried. The top panel is absolute USD, the bottom panel is inflation-adjusted USD.

Data for VA and CDMRP-funded projects (**Figures 4** and **5**) are not reflected by patient population, as they predominantly focus on military-relevant TBI in Service Members and Veterans. Similarly, NIDILLR grants (**Figure 6**) focused on TBI severe enough to cause long-term disability. In the graph depicting CDMRP funding, the first major increase occurred in 2007, coinciding with increases in deployment of US troops during Operation Iraqi Freedom/Operation Enduring Freedom (OIF/OEF). For the VA, the Translational Research Center for TBI and Stress Disorders (TRACTS) was initially funded in 2009 to examine imaging and other sequelae in Veterans with TBI. Large increases in funding from the CDMRP and the VA in 2012-13 and 2018-19 reflect, in part, the funding of the Chronic Effects of Neurotrauma Consortium (CENC) and subsequent Long-Term Impact of Military-Relevant Brain Injury Consortium (LIMBIC) projects. Other large projects include the Concussion Assessment, Research, and Education (CARE) Consortium and TBI Endpoints Development (TED) (Broglio et al., 2017; Manley et al., 2017). These three projects are described in the following section of this paper as well as further detailed in this issue.

**Figure 4.**
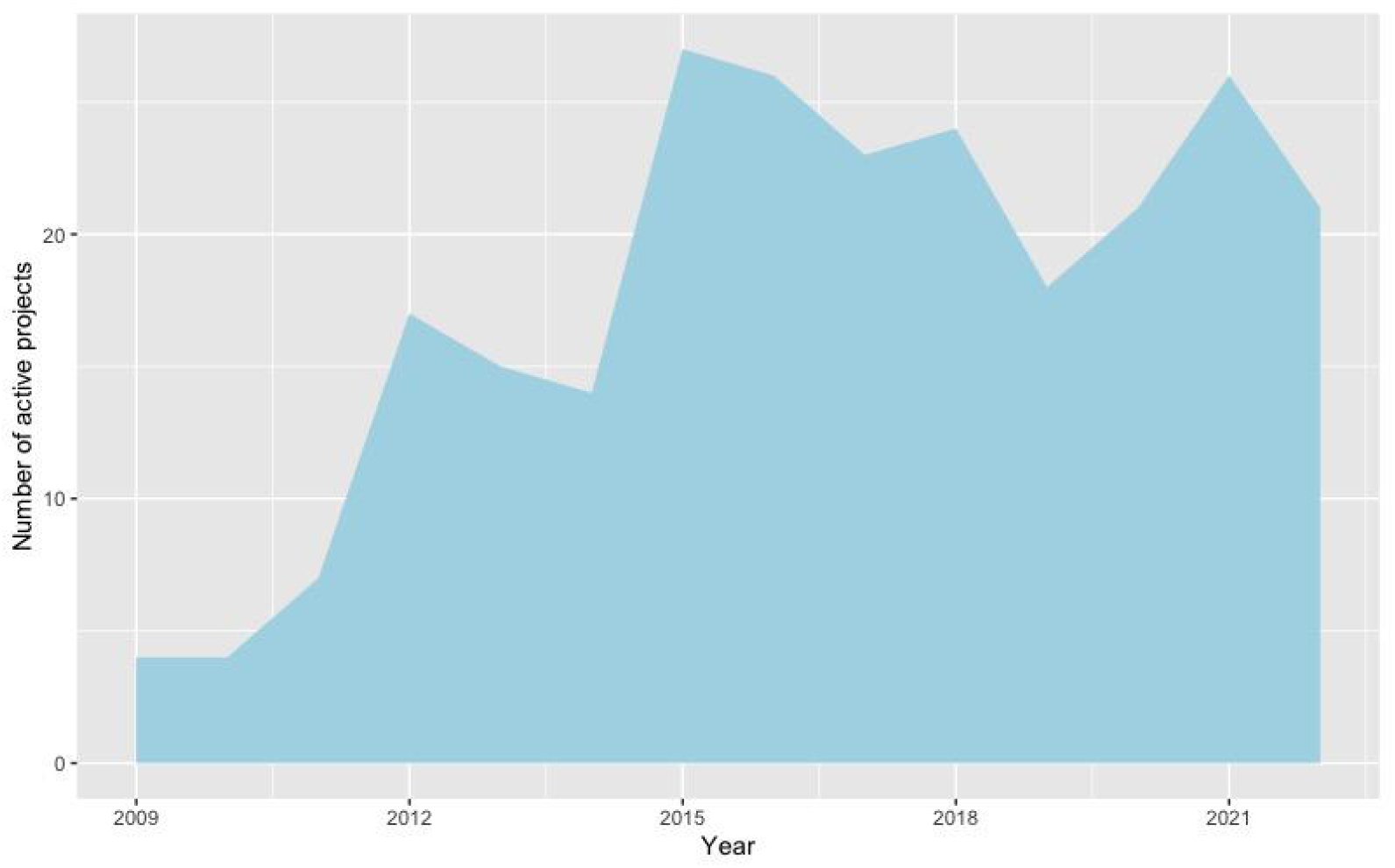
Veterans Administration (VA) funding trends. Graph depicts the number of active projects, rather than total funding, as NIH RePORTER does not list funding amounts for VA grants.

**Figure 5.**
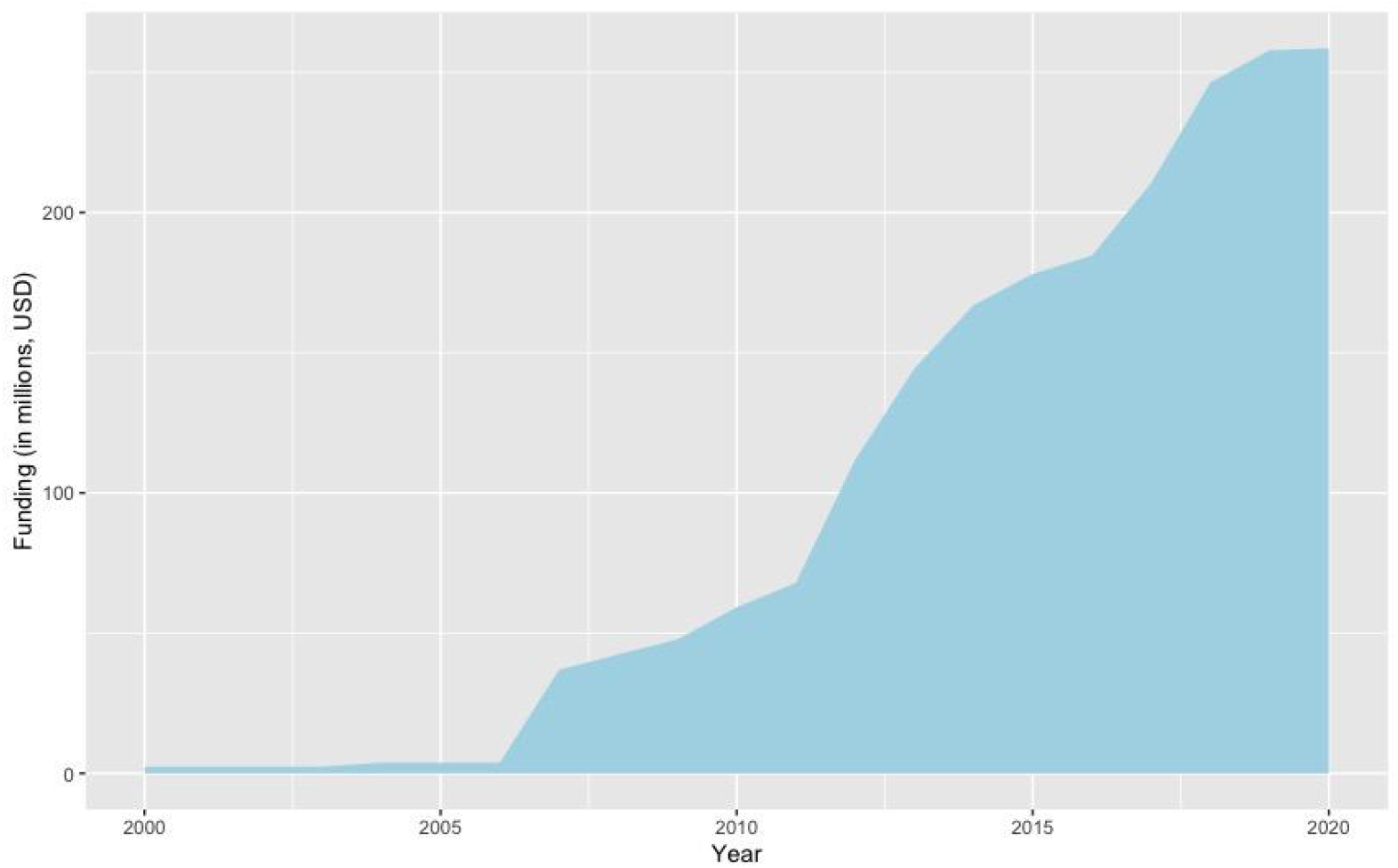
CDMRP funding trends. Amounts are shown in millions, in US Dollars. These amounts were only available for the full amount funded, without information on the duration of the project, so this graph depicts the *cumulative* funding committed from the first year of the project. The funding amount is not adjusted for inflation.

**Figure 6.**
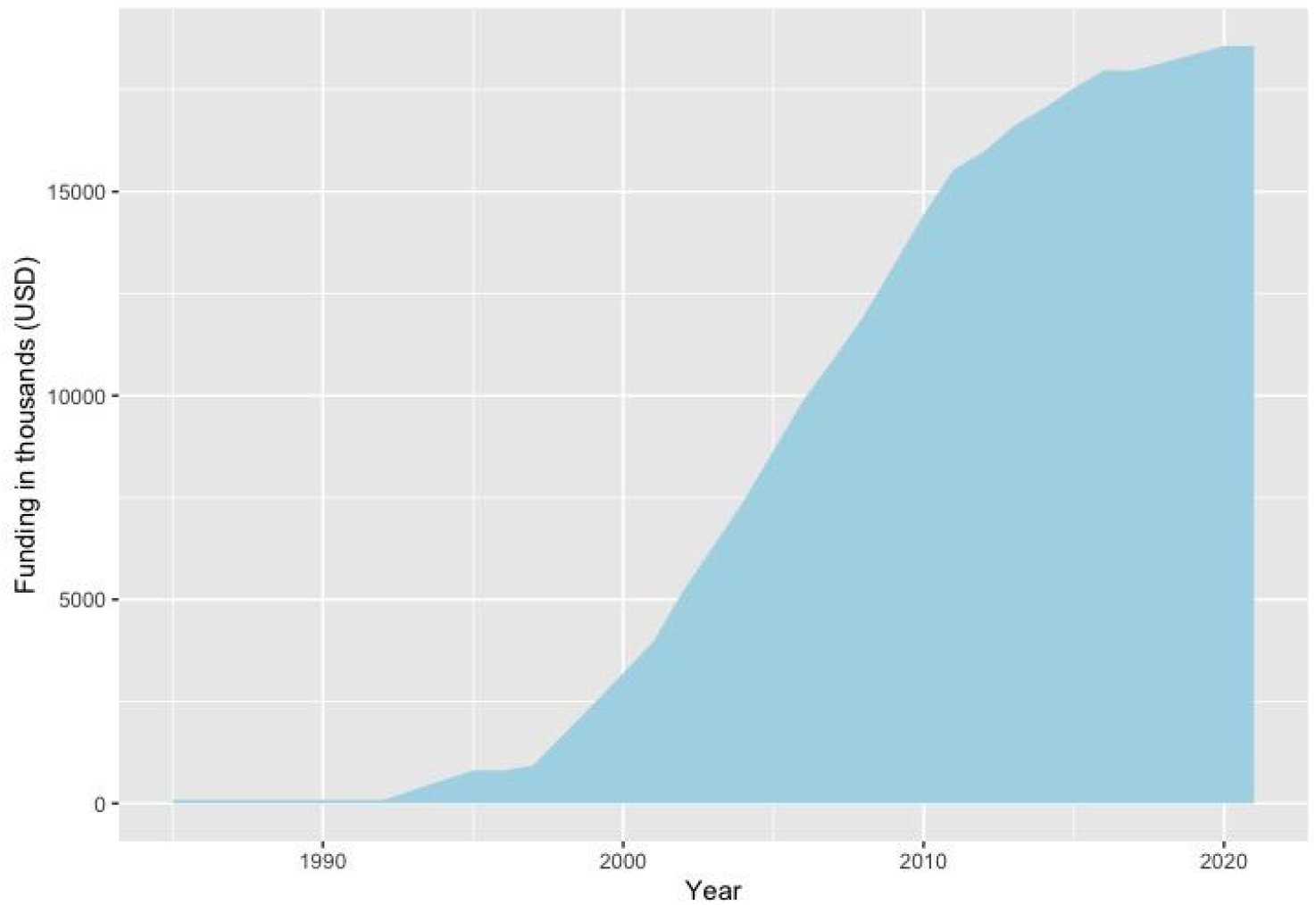
NIDILLR funding trends. NIDILLR grants focus on TBI severe enough to cause long-term disability. *Cumulative* funding is shown. Amounts are shown in thousands, in US Dollars.

There are some patient populations that are largely absent from these funding graphs, largely mirroring the trend in underrepresentation in the publication graphs.

Penetrating TBI often poses unique challenges for MR imaging owing to higher incidence of safety contraindications such as ferrous metal fragments as well as other debris which can create artifacts in imaging. Other challenges may relate to willingness of participants to engage in research or other conditions which may formally or practically exclude them from participation. Despite the prevalence of TBI and other head trauma in older adults, there are few funded projects focused on imaging this age group at any level of severity. This may reflect a significantly lower level of awareness in the general public, complexity of comorbid neurological and other health conditions in this age range, or other factors. However, recent funding by both NIH and VA have been awarded to extend TRACK-TBI objectives in older age ranges in both civilian and Veteran populations. Imaging studies of TBI occurring in the context of intimate partner violence (IPV) is also underrepresented, although a few recent studies have been funded to more closely examine this population with unique challenges (Esopenko et al., 2021).

### Large Multi-site Consortia

In both the US and other regions, there have been a number of multi-site consortia focused on TBI that have included neuroimaging data as a major component of the research. The extent and multimodal nature of these consortia allow for analysis on a larger scale and provide data that can begin to address the complexities of premorbid contributing factors and injury-relevant elements and sequelae.

#### Military-relevant mTBI

In 2013, the Chronic Effects of Neurotrauma Consortium (CENC) started collecting data at 8 sites across the U.S., including neuroimaging, blood samples, neurobehavioral, and neurocognitive assessments in OEF/OIF (Operation Enduring Freedom/Iraqi Freedom) Service Members and Veterans (Walker et al., 2016). This project was renewed and expanded in 2018 as Long-term Impact of Military-relevant Brain Injury Consortium (LIMBIC). The focus of the project is to examine the evolution of late effects of TBI (predominantly mild TBI) in U.S. Veterans and Service Members. Additional details regarding the efforts of this consortium are highlighted in a separate article in this issue.

#### Acute Emergency Department-based TBI

The Mission Connect Mild TBI Translational Research Consortium (2008-2013) collected observational data in patients with acute mTBI recruited from the emergency departments of Texas Medical Center-based hospitals. A subset of the participants participated in a clinical trial focused on statin intervention. Participants were enrolled within 24-48 hours post-injury and were regularly assessed up to 6 months post-injury. Data collected included neuropsychological assessments, blood samples for biomarkers, detailed clinical information, and neuroimaging data (including MRI and MEG data).

Started in 2013, The Transforming Research and Clinical Knowledge in Traumatic Brain Injury project (TRACK-TBI) began collecting clinical data, neuroimaging, blood biospecimens, and detailed outcomes in 3,000 patients from 18 Emergency Departments (EDs) in the United States across the injury spectrum (Yue et al., 2013). A similar project in the European Union started in 2013 as well, Collaborative European NeuroTrauma Effectiveness Research (CENTER-TBI), with similar biomarkers collected (Maas et al., 2015). The TBI Endpoints Development (TED) Initiative began in 2016 as a collaboration between groups of researchers to harmonize and curate the large datasets generated by larger clinical studies (Manley et al., 2017).

#### Sport-related Concussion

THE NCAA-DoD Concussion Assessment, Research, and Education (CARE) Consortium launched in 2014 to study student-athletes with neuroimaging across 6 sites, along with detailed assessments of head impact exposure, blood and genetic biomarkers, and neuropsychological outcome at multiple time-points (Broglio et al., 2017). Highlights of this consortium are featured in an article in this issue. The REPIMPACT Consortium, started in 2017, focused on adolescent soccer players, including neuroimaging, blood biomarkers, and neurobehavioral assessments at 6 sites across the European Union and Israel (Koerte et al., 2021).

#### Pediatric TBI

In 2017, Advancing Concussion Assessment in Pediatrics (A-CAP) began, focusing on children and adolescents presenting to the ED with mild TBI (mTBI) at 5 sites across Canada, including neuroimaging, psychosocial, and neurobehavioral measures (Yeates et al., 2017). In 2021, a new project started which focuses on identifying early predictors of post-concussive symptoms in adolescents, CARE4Kids. Assessments include neuroimaging, neuropsychological measures, blood markers, and autonomic functioning.

### Data Aggregation and Sharing Efforts

In addition to multi-site projects with coordinated data collection, initiatives focused on systematizing, aggregating, curating and sharing data have accelerated neuroimaging research in TBI.

#### Intra-Agency (NIH) Common Data Elements (CDEs)

The CDEs initiative, originally established in 2006 by the National Institute of Neurological Disorders and Stroke (NINDS) of the NIH was created to promote consistency and to facilitate data sharing through standardization of definitions, variables, protocols and structure for data collected in several neurological diseases. TBI was included in 2008 through a coordinated effort of multiple stakeholders funding TBI research, including the Department of Veterans Affairs (VA), the National Institute on Disability and Rehabilitation Research (NIDRR) of the Department of Education, the Defense Centers of Excellence for Psychological Health and Traumatic Brain Injury (DCoE) of the Department of Defense (DoD) and the Defense and Veterans Brain Injury Center (DVBIC) of the DoD. Recommended parameters were suggested for a limited set of imaging sequences, and common definitions and variables related to findings on CT and conventional MR sequences (e.g., T1-w, T2-w, FLAIR, SWI) were established (Duhaime et al., 2010; Haacke et al., 2010). These variables were later expanded in a subsequent version of the imaging CDEs to better incorporate findings more commonly identified in patients with mild and chronic injury as opposed to more acute and more severe injury (CDE Detailed Report | NINDS Common Data Elements). These CDEs have standardized coding of lesions and abnormalities in TBI on conventional imaging, enabling comparison across cohorts and facilitating larger studies than were previously possible.

### Federal Interagency TBI Research (FITBIR) Informatics System

The Federal Interagency TBI Research (FITBIR) informatics system (http://fitbir.nih.gov) is a secure, centralized database which serves as a central repository for new and previously collected data of different types (e.g., numeric, text, image, time series, etc.) using the specific variables developed as part of the TBI CDEs. FITBIR has been sponsored by the U.S. Army Medical Research and Materiel Command (funded through the Defense Medical Research and Development Program) and supported by the NIH Center for Information Technology, in addition to NINDS. In some circumstances, investigators may be required to submit data from federally-funded TBI-related projects to FITBIR, though submission of data from any relevant project, including older completed projects, is encouraged. Due to its size, FITBIR is also well poised to contribute to efforts in the field of TBI neuroimaging in consistency, data sharing and transparency.

#### Enhancing NeuroImaging Genetics through Meta-Analysis

The ENIGMA Consortium was created in 2009 (PI: Paul Thompson) with the goal of achieving adequate sample sizes to conduct well-powered genome-wide association studies (GWAS) with neuroimaging phenotypes (Thompson et al., 2020). It has since expanded to include numerous working groups focused on identifying reliable neuroimaging biomarkers for various psychiatric and neurological disorders (with or without genetic data). The ENIGMA Brain Injury working group was formed in 2016 and now includes multiple subgroups based on patient population or imaging modality (Wilde et al., 2019). ENIGMA is not a data repository, rather a collaboration framework, for which sharing raw data is optional. Coordinated processing and analysis, using meta- or mega-analyses, leads to sample sizes that may be large enough to apply big data approaches such as machine learning. There are also active projects on harmonization, including both neuroimaging data and cognitive data. Additional data on related to ENIGMA is highlighted in this issue (Esopenko et al., *this issue*)

### Data Harmonization

For more than a decade, funding agencies, publishers, and governmental agencies have required data management plans that address what has become known as FAIR data principles; Findable, Accessible, Interoperable, and Reusable (Wilkinson et al., 2016). This pushes researchers beyond the responsibility for proper data collection to support their own research questions, but to provide for the long-term care of the informational assets generated so that other researchers can “re-use” these data or combine them with new datasets. It is clear, however, that the infrastructure that has arisen from these efforts is not sufficient to ensure that archival data is usable in clinically meaningful ways dissociated from any sample biases or measurement errors that might impact analyses.

Relevant to this special issue on imaging, we have known for some time that there are often statistically significant differences in consortium level data acquired from different sites. This should not be unexpected as differences in vendors, equipment, and software versions can introduce a number of inconsistencies in the acquisition of imaging data. Early consortium studies like the Alzheimer’s Disease Neuroimaging Initiative (ADNI) invested huge amounts of time and money into trying to eliminate as many possible data compatibility issues by standardizing data acquisition between the sites, vendors, and investigators. These early attempts at “harmonization” entailed developing a set of very precise sequence settings for each vendor and software versions that was aimed at minimizing the many differences at the level of image acquisition. Many consortia continue to follow these guidelines by establishing sequence parameter settings for each site at the outset of the study so that acquisition is more consistent. However, even when this level of control of acquisition is successful, site differences can still persist.

Interests in aggregating legacy data only compound the issues impacting harmonization as the age of the study, differences in study design, study hypotheses, and the expertise of the groups acquiring the data lead to unique acquisition parameters and sequences. More recent efforts aimed at harmonizing data now aim to take existing datasets and use advanced mathematical procedures to limit the differences between sites or across studies. For example, one of the early methods developed to address differences in samples emerged in the genetics literature where microarray “batch” effects have been noted when samples were processed in separate experiments within samples. Small differences between these batches make it inappropriate to combine these data without adjusting for these differences. In 2007, Bayes statistical methods were first proposed as a statistical approach for accounting for these batch differences with moderate success (Johnson et al., 2007). This method became known as ComBat (COMbining BATches) and it is becoming widely used in neuroimaging research when existing processed imaging results are available. Currently, there are numerous peer reviewed reports and large consortia (Radua et al., 2020) that use these methods (or modified versions of this method) to improve the compatibility of neuroimaging data, ultimately improving the sensitivity of aggregated data to capture important clinical features.

There are also emerging methods for harmonizing imaging data at the raw imaging data level. This includes efforts to collect the raw unprocessed k-space data as well as applying methods to the exported DICOM data. Raw unprocessed k-space data has the advantage of being unprocessed by proprietary vendor specific software. In its raw form, it can be pre-processed in a consistent manner that then eliminates potential vendor specific quantification variability. However, the size of the imaging data represents a significant limitation for some. Methods that use the DICOM data can also be used where the images can be that essentially “rewrites” the data as an image file that accounts for the differences in spatial signal variability at different sites (Cetin Karayumak et al., 2019; Mirzaalian et al., 2016, 2015).

In summary, there are a growing number of options that can be used to address the need for image harmonization. As these methods grow, the FAIR principles can be practically applied to existing data well beyond the infrastructures built to support the storage and curation of publicly funded data sets.

### Multimodality Integration Efforts

Multimodal data collection is hardly new, with the first multimodal paper in our review being published in 1987, but it has represented a growing portion of the literature, with 25-30% of papers in recent years including more than one neuroimaging modality. Collecting multimodal data is not the same as *integrating* multimodal data, however. At the most basic level, integrating multimodal data could be simply using multiple measures jointly in a prediction of outcome, as they may each contribute unique information (Finitzo et al., 1987). In more recent years, one common way of using multimodal data involves creating connectivity networks representing the structural or functional connections in the brain. This requires a standard anatomical scan such as a T1-weighted MRI along with a sequence that maps the white matter tracts in the brain (diffusion MRI) or one that can be used to show correlations in brain activity over a period of time (resting state fMRI) (Caeyenberghs et al., 2017, 2012; Dennis et al., 2017; Han et al., 2014). Another step in multimodal integration is integrated data processing, with co-registration of two or more modalities and summary metrics extracted from common regions (Dennis et al., 2018). Lastly, some techniques involve statistical integration of data using advanced machine learning and statistical methods, which allow for multi-dimensional examination of data (Avants et al., 2021; Zavaliangos-Petropulu et al., 2017).

### Future Directions in the Translation of Imaging in TBI Treatment

#### Normative data

One of the primary challenges of translating neuroimaging findings to treatment at the level of the individual patient is the relative lack of normative data to generate standards. Research studies generally use case/control models and group comparisons, but the field generally lacks the very large amounts of well-characterized, healthy controls necessary to create normative data for quantitative MRI methods. There are current efforts to create these cohorts, including components of TRACK-TBI and ENIGMA as well as the development of the Neuroimaging Normative Library project (https://www.cohenveteransbioscience.org/programs/our-programs/normative-neuroimaging-library/). Additionally, tools like https://centilebrain.org/#/ that allow individual sites to calculate standard scores for subcortical and cortical measures based on data from thousands of individuals around the world.

#### Pipeline optimization

Several groups of investigators are also engaged in efforts to evaluate the relative advantages and challenges of different existing pipelines related to imaging analysis in TBI and other conditions, refine existing analytic methods and create new tools, as necessary. Such projects are embedded in the central objectives of some of the larger consortia efforts, including LIMBIC and ENIGMA, and endeavor to optimize methods and quality control procedures as well as to enhance consistency, accessibility, usability, and replicability across groups. In addition to efforts to develop analysis methods, efforts to provide training materials and in person or webconference training in their use may accelerate the pace of research in this area. Additionally, enhanced accuracy and speed of analysis are necessary for eventual application in clinical practice.

### Conclusion

TBI neuroimaging research over the past 55 years has produced a vast amount of data that has greatly increased our understanding of injury and recovery processes. However, many types of imaging, particularly more recent or advanced modalities, have not yet been implemented in clinical practice. This is not a reflection of the quality of work that has been done by the field to date, but is primarily due to patient heterogeneity complicating efforts to identify reliable imaging biomarkers. Large sample sizes, either through prospectively harmonized joint data collection efforts or through retrospectively sharing and harmonizing, will further our ability to generate predictive models for individual patients. The data, the expertise, and the techniques exist, but optimizing and integrating these will hasten discovery and advance clinical utility.

## Data Availability

Data are all from public databases

## Acknowledgements

ELD, FK, DFT, and EAW are supported by NIH/NINDS R61NS20249 and R01NS122184 and by VA I01RX0000341144.

## Supplementary Materials

**Supplementary Notes 1**. PubMed Search Terms

**Supplementary Notes 2**. NIH RePORTER Search Terms

**Supplementary Notes 3**. CDMRP Search Terms

**Supplementary Notes 1**. PubMed Search Terms

((TBI[Title/Abstract] OR traumatic brain injury[Title/Abstract] OR head injury[Title/Abstract]) NOT (pig[Title/Abstract] OR rat[Title/Abstract] OR mouse[Title/Abstract] OR porcine[Title/Abstract])) AND (MR[Title/Abstract] OR magnetic resonance [Title/Abstract] OR PET[Title/Abstract] OR positron[Title/Abstract] OR neuroimaging[Title/Abstract] OR ASL[Title/Abstract] OR QSM[Title/Abstract] OR SWI[Title/Abstract] OR arterial spin labeling[Title/Abstract] OR susceptibility[Title/Abstract] OR MEG[Title/Abstract] OR EEG[Title/Abstract] OR electroencephalo[Title/Abstract]))

**Supplementary Notes 2**. NIH RePORTER Search Terms.

Fiscal Year: Active Projects, 2021, 2020, 2019, 2018, 2017, 2016, 2015, 2014, 2013, 2012, 2011, 2010, 2009, 2008, 2007, 2006, 2005, 2004, 2003, 2002, 2001, 2000, 1999, 1998, 1997, 1996, 1995, 1994, 1993, 1992, 1991, 1990, 1989, 1988, 1987, 1986, 1985

Text Search: (TBI or traumatic brain injury or concussion or head injury or concussive) and (MRI or (magnetic and resonance) or PET or positron or neuroimaging or ASL or QSM or SWI or DTI or DWI or (arterial and spin and labeling) or MEG or EEG or magnetoenceph or electroenceph or (diffusion and imaging)) (advanced)

Limit to: Project Title, Project Terms, Project Abstracts Combine multiple categories with: OR NIH Spending Category (not used for VA grants): Biomedical Imaging, Injury - Childhood Injuries, Injury - Trauma - (Head and Spine), Injury - Traumatic brain injury, Injury - Unintentional Childhood Injury

**Supplementary Notes 3**. CDMRP Search Terms

Fiscal Year: All Fiscal Years

Research Program: ARIF, DMRDP, DRMRP, JWMRP, PH-TBI, PRARP, PRMRP

Abstract Keywords: TBI OR brain injury OR concussion

**Supplementary Figure 1.**
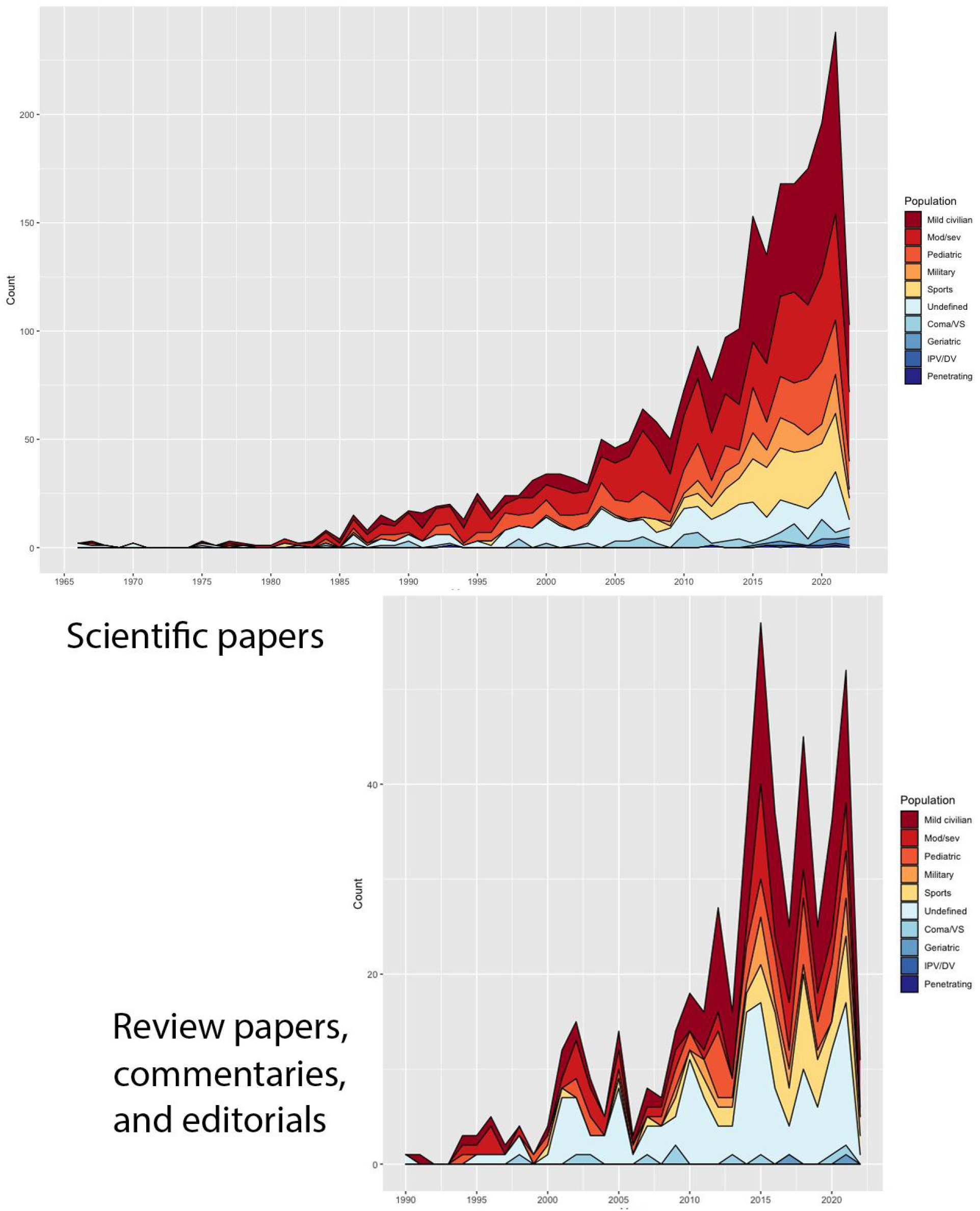
Publication trends classified by patient population with scientific papers separated from review/editorial/commentary papers. VS=vegetative state, IPV/DV=intimate partner violence/domestic violence, mod/sev=moderate/severe.

**Supplementary Figure 2.**
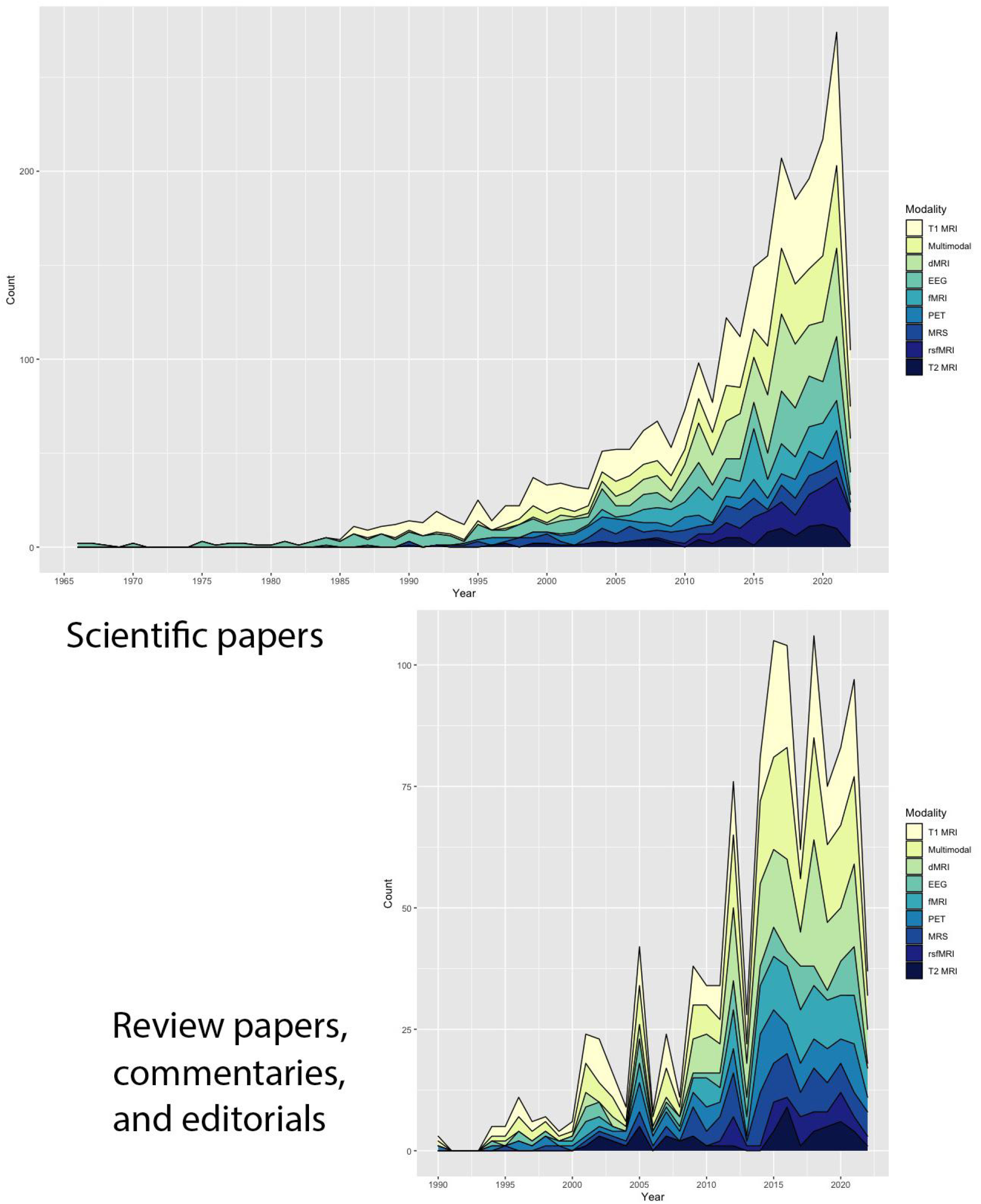
Publication trends classified by modality with scientific papers separated from review/editorial/commentary papers, part 1. Data are split into two panels for easier viewing. dMRI=diffusion magnetic resonance imaging, EEG=electroencephalography, fMRI=functional MRI, MRS=magnetic resonance spectroscopy, PET=positron emission tomography, rsfMRI=resting state fMRI.

**Supplementary Figure 3.**
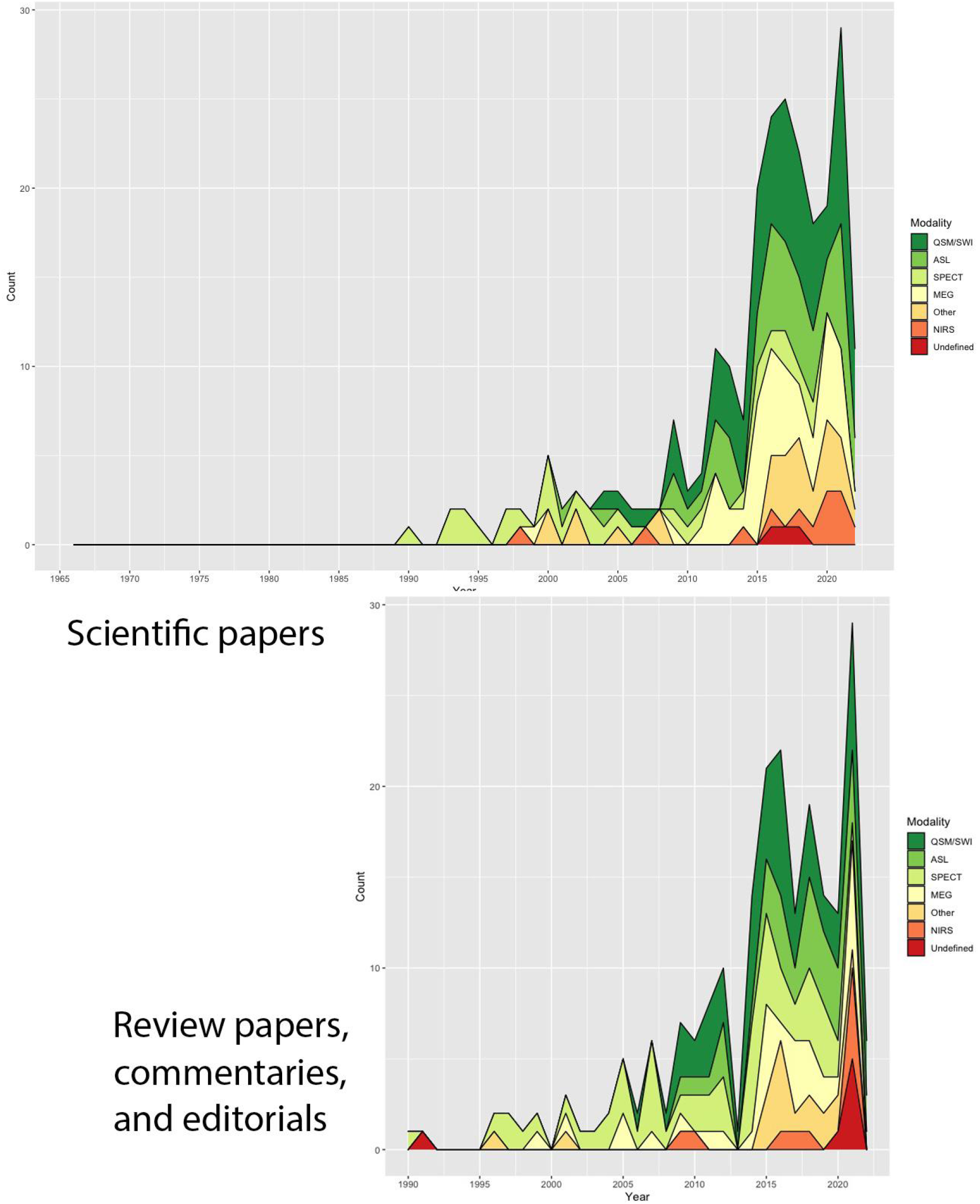
Publication trends classified by modality with scientific papers separated from review/editorial/commentary papers, part 2. Data are split into two panels for easier viewing. ASL=arterial spin labeling, MEG=magnetoencephalography, NIRS=near-infrared spectroscopy, QSM=quantitative susceptibility mapping, SWI=susceptibility weighted imaging, SPECT=single photon emission computed tomography.

